# Improved cognition after high-intensity exercise paired with motor practice in individuals with stroke and older adults: A randomized controlled trial

**DOI:** 10.1101/2023.02.09.23285669

**Authors:** Brian Greeley, Beverley C. Larssen, Jennifer Ferris, Negin Motamed Yeganeh, Justin W. Andrushko, Briana Chau, Christina B. Jones, Sue Peters, Kristin L. Campbell, Jason L. Neva, Lara A. Boyd

**Affiliations:** Department of Physical Therapy, University of British Columbia, Vancouver, Canada; Clinical Research, Surrey Memorial Hospital, Fraser Health, Surrey, British Columbia, Canada; Graduate Program in Rehabilitation Sciences, University of British Columbia, Vancouver, Canada; Gerontology Research Centre, Simon Fraser University, Vancouver, Canada; Djavad Mowafaghian Centre for Brain Health, University of British Columbia, Vancouver, Canada; School of Physical Therapy, University of Western Ontario, London, Canada; Université de Montréal, École de kinésiologie et des sciences de l’activité physique, Faculté de médecine, Montréal, QC, Canada, H3C3T5; Centre de recherche de l’institut universitaire de gériatrie de Montréal, Montréal, QC, Canada, H3T1P1

**Author notes:** Corresponding author: Lara Boyd, PT PhD, Department of Physical Therapy University of British Columbia Vancouver, Canada V6T 1Z4. Financial interests: The authors declare no competing financial interests.

**Keywords:** Chronic stroke, high-intensity interval training, exercise, transfer, cognition, motor practice

## Abstract

**Introduction:** Stroke is a leading cause of long-term disability resulting in cognitive and motor impairments. Exercise may improve cognition and motor function. We paired multiple bouts of high-intensity interval training (HIIT) exercise with motor practice to positively affect cognitive and motor function after stroke and age-matched controls.

**Methods:** Using a randomized controlled parallel group design, 31 individuals with chronic stroke and 41 older adult controls were randomized to either 23 minutes of HIIT exercise or rest prior to completing motor task practice using their paretic/non-dominant arm across five days. Primary outcomes were reaction time and motor function. Assessors were blinded to the intervention group. Trail Making Test-A and B (TMT-A, TMT-B), and object hit and avoid (OHA) were used to assess processing speed and inhibitory control.

**Results:** All participants showed evidence of motor learning; HIIT exercise did not confer an additional benefit. For stroke participants, motor function (*p* = .047), but not motor impairment, improved. The stroke exercise group displayed significant reductions in TMT-A completion time (*p* = .026). Exercise with motor practice also led to a reduced number of distractors hit (*p =* .035) in the OHA task for both participant groups. There were no adverse events.

**Conclusions:** Five days of HIIT exercise paired with motor practice led to improved processing speed for individuals with stroke. Both participant, exercise groups showed improved visuospatial skills and inhibitory control. Together, HIIT exercise paired with motor practice appears to be a safe and effective means of enhancing cognitive-motor skills after stroke and in older adults.

**Trial Registry:** ClinicalTrials.gov, ID: NCT02980796

**Trial registry name:** The Influence of Exercise on Neuroplasticity and Motor Learning After Stroke (EX-ML3)

## Introduction

Stroke is the leading neurological cause of adult physical disability (Cieza et al., 2020), responsible for cognitive and motor impairments. Fifty-two percent of those who have motor recovery continue to show signs of cognitive impairment three years after stroke (Kapoor et al., 2017). Identifying effective interventions that target recovery of both motor and cognitive functions is critical to improving the quality of life after stroke.

Exercise delivered as a standalone intervention appears not to impact cognition after stroke. For example, a single treadmill walking session at 70% of heart rate reserve resulted in an improvement in upper limb function, but did not benefit cognition in individuals in the chronic phase of stroke recovery (Ploughman et al., 2008). Similarly, a multi-session aerobic exercise intervention led to improved paretic hand function without affecting cognition in individuals with chronic stroke (Quaney et al., 2009). While a meta-analysis including only randomized controlled trials found that physical activity had a positive impact on cognition after stroke, this effect disappeared when including studies that only used aerobic-only interventions (Oberlin et al., 2017). On the other hand, cognitive function may improve by pairing exercise with practice of a demanding cognitive task. Increased fluid intelligence was noted in individuals with chronic stroke following 10 weeks of pairing aerobic exercise with adaptive cognitive training (Ploughman et al., 2019). While the findings from these studies suggest that exercise may boost the effects of cognitive training, it is unknown whether exercise paired with a motor task would transfer to untrained motor tasks that require high cognitive demand (i.e., cognitive-motor tasks).

In the current study, we paired high-intensity interval training (HIIT) exercise, given its effectiveness to promote cognitive plasticity (Hugues et al., 2021), its feasibility, and safety in those who have had a stroke (Boyne et al., 2013; Charalambous et al., 2018; Gjellesvik et al., 2020), with practice of a motor task in a group of individuals who were in the chronic phase of recovery from stroke, and a group of healthy older adults. Participants underwent 5 separate sessions consisting of 23 minutes of either HIIT exercise or rest prior to motor practice. Motor learning, function and performance on cognitive-motor tasks were assessed pre- and post-intervention. We hypothesized that individuals in the exercise group would show enhanced motor learning and function, similar to previous findings in stroke (Quaney et al., 2009). We also hypothesized that the exercise group would show cognitive transfer, or an improved ability to perform untrained, cognitively engaging motor tasks (Ploughman et al., 2019), relative to the rest group.

## Methods

### Participants

We used past work that employed a HITT exercise intervention paired with motor practice in two groups (Mang et al., 2014) and G*Power to calculate power. Motor behaviour at retention (116 ms for rest, 98 ms for exercise) and a common standard deviation (sigma = 22.6) with an α = 0.05 and power of 0.80 (two-samples), showed that 26 participants per group or a total of 52 individuals with stroke and an equal number of matched controls, were necessary. Fifty-five participants in the chronic phase of stroke recovery (>6 months from stroke incident) and fifty older adults were screened for eligibility to participate, with 40 stroke participants and 46 controls consenting to participate in the study (Figure 1). Note, recruitment numbers were lower than planned given data recruitment disruptions and costs owing to mandatory COVID related research suspension. Participants were considered eligible if they were between the age of 40-85, left or right hand dominant, and showed no signs of cognitive impairment (Milani et al., 2018) as assessed by the Montreal Cognitive Assessment (Nasreddine et al., 2005). Exclusion criteria included: diagnosed with neurological or psychiatric conditions other than stroke, comprehensive aphasia, contraindications to exercise, failure to see targets, or inability to extend and maintain the paretic arm/hand inside a target for 500 ms on a Kinarm end-point robot (Kinarm, Kingston, Ontario).

**Figure 1.**
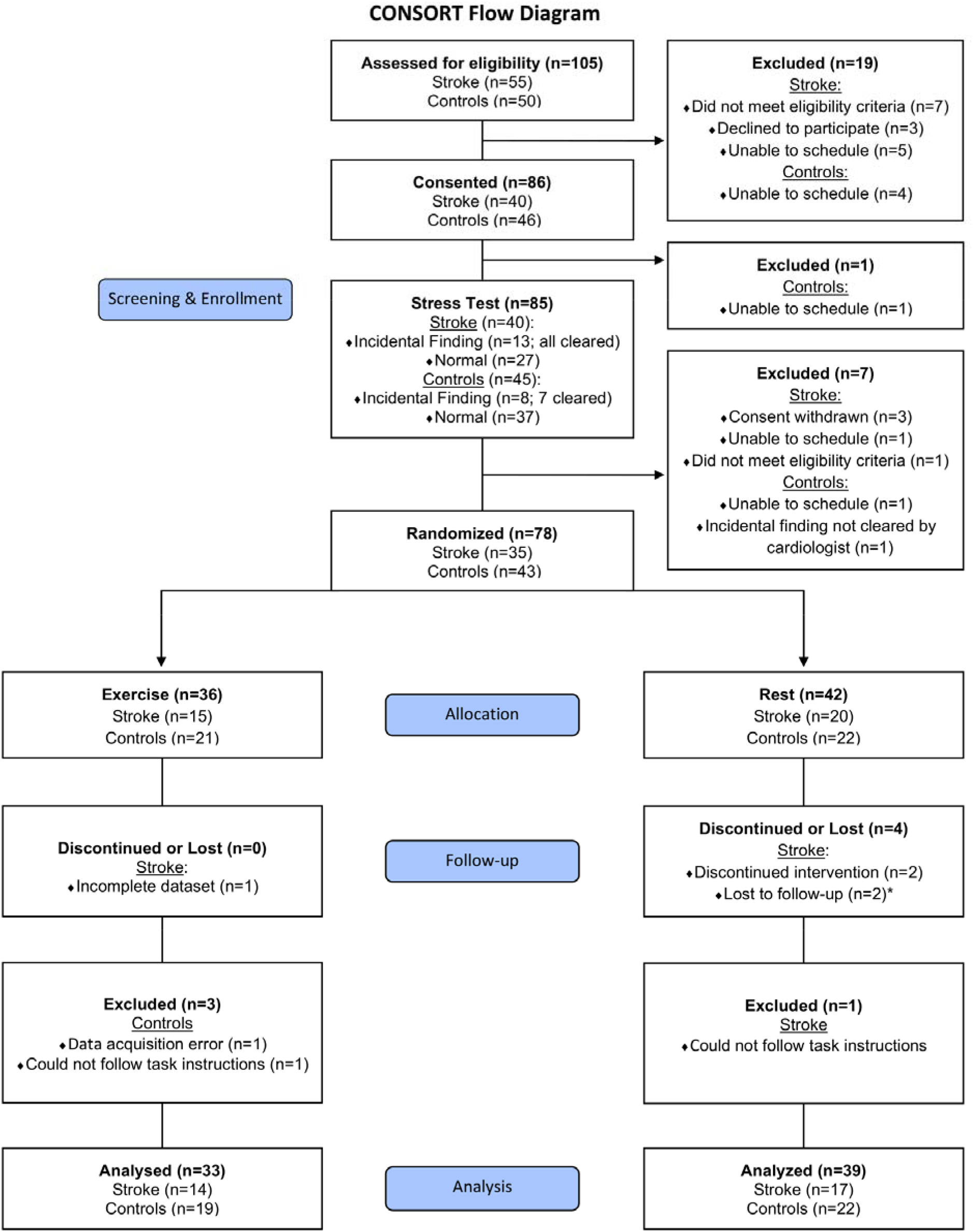
Consolidated standards of reporting trials (CONSORT) flow chart. * denotes the 2 participants’ 35-day retention data were missing due to the COVID-19 research closure. Data from these 2 individuals were retained for analysis when possible.

This study was approved by the University of British Columbia Ethics Committee (Clinical Research Ethics Board #H16-01945); all participants read and signed a consent form prior to any experimental protocols. All data collection was performed at the University of British Columbia Hospital. Subsets of these data have been previously published (Andrushko et al., 2023; Greeley et al., 2021; Neva et al., 2022); however, the questions and analyses and data subset in the present manuscript are novel.

### Study design

This parallel group randomized controlled trial consisted of 11 sessions (Figure 2). A cardiologist supervised maximal stress test cleared individuals for exercise (session 1). Following the stress test, participants were pseudorandomly allocated into either HIIT exercise or rest groups. Randomization was completed using a custom Matlab script stratifying participants based on sex, age, and motor impairment (if stroke). Our target allocation ratio was 1:1 across groups and experimental conditions. One study team member not involved in data collection, completed the randomization, and communicated the intervention allocation to experimenters. Study participants nor experimenters were blinded to group assignment; however, clinical assessors were blinded to group. Across 5-days (sessions 4-8), participants engaged in 23 minutes of either HIIT exercise or watched 23 minutes of a nature documentary immediately before practice of a motor task. Upper-extremity impairment and function, and cognitive assessments were conducted pre- (session 2) and post-intervention (session 9 at a 24-hour, short-term retention test). Participants returned for a second retention test (session 11) between 30-35 days later to assess long-term change associated with motor learning. Sessions 3, 7 and 10 contained neurophysiological assessments and are not reported here.

**Figure 2.**
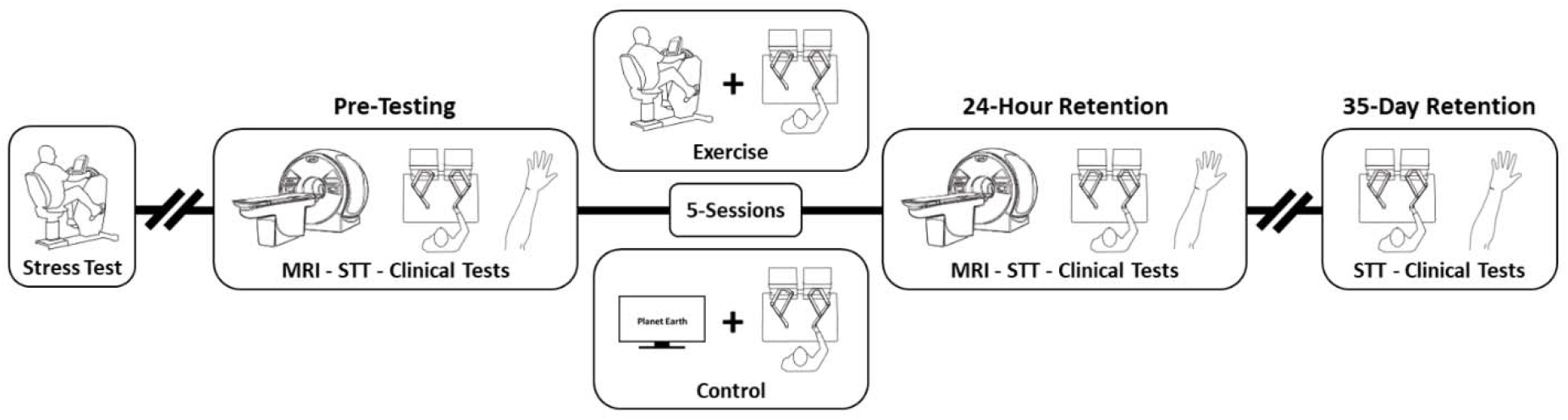
Study design. On day 1, all participants underwent a stress test. On day 2 and 8, all participants underwent magnetic resonance imaging scans and completed the Kinarm standard test battery, whereas only stroke participants also completed Fugl-Meyer and the Wolf Motor Function Test to test upper limb impairment and function, respectively. Kinarm test battery included Trail Making Test A & B and object hit and avoid, two tasks used to assess motor-cognitive transfer. On day 4-8, participants completed 23 minutes of exercise or rest followed by 4 blocks of paretic/non-dominant upper-limb motor practice of the serial targeting task, an implicit motor sequence task, on the Kinarm. On day 9, 24 hours after day 8, participants completed 1 block of the serial targeting task followed by Trail Making Test A & B and object hit and avoid. On day 11, 35 days after the last practice day (i.e., day 8), participants completed 1 block of the serial targeting task.

### Stress Test

To ensure safety, assign individualized intensity during exercise sessions, and best practices (MacKay-Lyons et al., 2020), all participants completed a physician supervised maximally graded stress test on a recumbent bike (SCIFIT, Tulsa, OK, USA). After 3 minutes of rest, baseline resting heart rate (HR) and blood pressure (BP) were recorded. Participants were then seated on a recumbent bike, identical to the one used in sessions 4-8. HR and rating of perceived exertion (RPE)(Heath, 1998) were monitored and recorded at every minute. BP was assessed every 2 minutes by a cardiology technologist. Participants were instructed to keep the pedaling cadence between 50-80 revolutions per minute (RPM). The stress test began with a 2-minute warm-up at 10 Watts (W), after which resistance increased either by 5, 10, or 15W based on current stress test recommendations (Beltz et al., 2016). Resistance was increased until either the participant reached volitional exhaustion, or RPM dropped below 50 and continued to decline for 5 seconds. After the stress test, participants cooled down for 3 minutes at 10W, then rested until HR and BP returned to baseline.

### Intervention

#### Exercise and rest

During each practice session, participants in the exercise group completed a 5-minute warm-up, then 3 × 3-minute intervals of high-intensity exercise (75% of their maximum wattage achieved during the maximal stress test) with a 3-minute active recovery (10 W) for a total of 23 minutes on a recumbent bike. This method was chosen based on its availability in rehabilitation settings and its safety and feasibility in stroke participants (Charalambous et al., 2018). Participants pedaled between 50-80 RPM throughout each session. HR was monitored from the non-paretic wrist with an Alpha 53p heart rate watch (Mio, Portland), BP and RPE were taken in the last 30-60 seconds of each interval. Participants in the rest group watched a nature documentary on a 50-inch screen for 23 minutes. HR was monitored every 3 minutes.

#### Motor task practice: Serial Targeting Task

After exercise or rest, all participants practiced 4 blocks of the Serial Targeting Task (STT), an implicit motor sequence task performed on a Kinarm End-Point robot (Greeley et al., 2021). Individuals were seated in a Kinarm chair, adjusted so that the participant’s head was positioned in the center of the visual field. Participants used their paretic/non-dominant upper-limb (visually occluded by a bib) to practice 4 blocks of the STT, over 5-days (2,220 total movements). Participants were instructed to move their hand to each target as quickly and accurately as possible.

The STT had a total of 9 possible targets. Eight targets formed an equidistant circular array around the 9th target (Supplementary Figure 1). Targets were displayed on the participant’s paretic/non-dominant visual field. Only one target was visible at any given time; to initiate the appearance of the next target, participants were required to hold the cursor within the current target for 500 ms. Participants had 10,000 ms to reach the target. For the pre-test, participants performed 20 target reaches. Motor acquisition was assessed each practice day whereas motor learning was assessed 24-hours and 35-days following the last day of practice.

Unknown to participants (see Supplementary Methods: Explicit Awareness), there was a repeating 6-element sequence (Supplementary Figure 1C) flanked by a random sequence of 7 targets. Random sequences appeared in the same order for all participants but did not follow a pattern or ever repeat. Participants were exposed to 32 repetitions of the repeating sequence (8 times per block, 4 blocks of practice; 192 trials) and to 36 random sequences within each practice session.

Reaction time (RT) was used to characterize change in motor behaviour; RT was averaged for each individual sequence trial, then calculated as a ratio relative to the averaged pre-test for the corresponding trial sequence type (e.g., averaged pre-test random trial / averaged first random trial in practice). Thus, accounting for potential differences in initial RTs across individuals and groups. Ratios were calculated for each practice day. Higher ratios indicated faster RT relative to the pre-test. Change in ratios for the random sequences indicate alterations in performance while changes in repeated sequences ratios indicates learning. Change in RT relative to pretest was a pre-specified primary outcome used to index motor learning.

### Assessments

#### Kinarm standard test battery

In the pre- and post-intervention sessions, the Kinarm was used to assess Trail Making Test-A and B (TMT-A, TMT-B), and object hit and avoid (OHA) for all participants. These are a part of the Kinarm Standard Tests™ battery and were collected as additional outcome measures of interest.

#### Trail Making Test A and B

TMT-A and TMT-B were used to assess processing speed and task switching, respectively (Vasquez & Zakzanis, 2015). To complete the task, participants grasped and held one of the robotic arms with their non-paretic/dominant hand. Hand position was represented by a small, solid white dot. Participants were asked to connect 25 numbered (TMT-A), or 13 numbered and 12 lettered circles (TMT-B) in ascending order. If participants made an error by moving to an incorrect target, the last correct target turned red; participants were required to return to the red target to resume the task. All stimuli were presented on the non-paretic/dominant side of the Kinarm workspace. TMT-A was always completed before TMT-B. Total completion time (i.e., the time in seconds from task onset to when participants touched the last target) was the dependent variable for both assessments.

#### Object Hit and Avoid

OHA is a visuomotor task that assesses rapid bimanual motor decisions, attention, and inhibitory control (Bourke et al., 2016). As the participants observed the two red targets (e.g., circle, rectangle) displayed on the Kinarm workspace, they were instructed to use two visually displayed 5 cm wide green paddles as their hands to hit away as many of these targets as possible. During the task, distractor targets were also present (i.e., oval, square) and participants were instructed to avoid hitting them. The speed and number of moving stimuli increased as the task progressed such that a single, slow (~10 cm/s) stimulus was visible at the beginning of the task and a maximum of 16 fast (~50 cm/s) stimuli were present at the end of the task. A total of 300 stimuli (200 targets, 100 distractors) were randomly presented in approximately 2.5 minutes. The number of target hits and distractor hits were calculated separately for each hand and used as the dependent variables.

#### Upper-extremity motor function and impairment in stroke

Trained clinical assessors administered and scored the Wolf Motor Function Test (WMFT) and the Fugl-Meyer Assessment (FMA) pre- and post-intervention. The WMFT indexes arm motor function and contains 15 timed movement tasks. If no repetitions were completed within 120 seconds for a task, a zero score was assigned. Each task was characterized via calculation of rate (repetitions/60 second); higher rates show faster movements and better motor function (Hodics et al., 2012). WMFT rate was the pre-specified primary outcome collected to characterize potential changes in motor function as a result of the intervention. The upper extremity portion of the FMA (/66) was collected as an additional outcome of interest to characterize paretic arm impairment, with higher scores reflecting less impairment (Lin et al., 2004).

## Statistical Analysis

All statistical analyses were carried out using SPSS software (SPSS 27.0; IBM Corporation, Armonk, NY). Data were tested for normality with the Shapiro-Wilk test with α = .001 (Gamst et al., 2008). When skewed or kurtotic, we performed the analysis on log-transformed data, however, non-transformed data are presented in the figures.

Independent samples *t*-tests were used to compare baseline demographic data between exercise and rest subgroups within stroke. We performed mixed repeated measures analysis of variance (RM-ANOVA) for all outcome measures. For our primary outcome, RT data from the STT, we ran a 5 (Practice Day: 1,2,3,4,5) × 2 (Sequence: repeated, random) × 2 (Intervention Group: rest, exercise) × 2 (Participant Group: stroke, older adults) RM-ANOVA to test motor acquisition. We also performed a 2 (Retention Day: 24-hour, 35-day) × 2 (Sequence: repeated, random) × 2 (Intervention Group: rest, exercise) × 2 (Participant Group: stroke, older adults) RM-ANOVA test on the RT data from the STT to assess motor learning. For OHA, a 2 (Hand: paretic, non-paretic) × 2 (Session: pre-, post-intervention) × 2 (Intervention Group: rest, exercise) × 2 (Participant Group: stroke, control) RM-ANOVA was run. To ensure changes in number of targets or distractor targets were not due to a speed-accuracy trade-off, we also completed a separate RM-ANOVA on hand speed in the OHA (see Supplementary Results). Multiple one-way RM-ANOVA tests were used on all single hand outcome measures (WMFT, FMA, TMT-A, TMT-B). Each participant’s physical fitness (blood pressure: diastolic and systolic at rest, resting HR, Godin Leisure-Time Exercise Questionnaire (Godin, 2011), and max watts achieved during the stress test) was calculated as a composite z-score and used as a covariate in all analyses (Greeley et al., 2021). We employed Šídák corrections for multiple comparisons (α < .05), and partial eta squared (η_p_^2^) effect sizes are reported.

## Results

Eight-six participants were enrolled between May 2017 and January 2022. Participant recruitment was paused between March 2020 and July 2021 due to COIVD-19. After exclusion, 31 individuals with stroke (mean age = 68.48 ± 11.18; n = 14 exercise; n = 17 rest) and 41 healthy older adults (mean age = 66.24 ± 7.91; n = 19 exercise; n = 22 rest) were included in the final analyses as per their original group allocation (Figure 1; see Supplementary Results: Participant Dropout). There were no adverse events associated with exercise testing or training.

### Stroke Baseline Demographics

There was a group difference in the maximum HR during the stress test with those in the exercise group displaying a greater HR relative to the rest group (*p* = .047). However, it should be noted that 6 participants in the rest group were on beta-blockers as opposed to 2 in the exercise group. There were no other differences in baseline demographics across groups (all *p*’s > .227). See Table 1 for complete summary of stroke participants demographics.

**Table 1.** Demographics of the stroke participant group. BB = Beta-blocker; Godin = Godin Leisure-Time Exercise questionnaire; FM = Fugl-Meyer; HR = heart rate; MOCA = Montreal Cognitive Assessment; TSS = time since stroke (months); WMFT = Wolf Motor Function Test. * = participant not included in secondary analyses; ¥ = participant has expressive aphasia; # = did not return; ^ = missing.

| Rest | ID | Sex | TSS | Endinburgh | Affected Arm | Pre FM | 24-h FM | Pre WOLF | 24-h WOLF | HR | Max HR | Max Watts | Godin | MOCA | BB Use |
| --- | --- | --- | --- | --- | --- | --- | --- | --- | --- | --- | --- | --- | --- | --- | --- |
|  | 1 | M | 54 | 50 | R | 64 | 61 | 73 | 64 | 71 | 157 | 120 | 97 | 27 | N |
|  | 2 | M | 84 | 60 | L | 55 | 61 | 43 | 50 | 78 | 104 | 60 | 60 | ^ | N |
|  | 3 | M | 110 | 70 | R | 63 | 62 | 73 | 47 | 84 | 108 | 50 | 14 | 27 | Y |
|  | 4 | M | 51 | 0 | R | 57 | 44 | 30 | 30 | 58 | 84 | 35 | 31 | 22 | Y |
|  | 5 | F | 45 | 79 | R | 60 | 59 | 39 | 37 | 75 | 157 | 40 | 65 | 28 | N |
|  | 6 | M | 16 | 70 | L | 63 | 60 | 49 | 54 | 45 | 113 | 115 | 15 | 25 | N |
|  | 7 | F | 228 | ^ | R | 28 | 27 | 15 | 15 | 65 | 102 | 25 | 15 | ^ | N |
|  | 8 | F | 143 | 90 | L | 62 | 63 | 48 | 52 | 54 | 100 | 65 | 24 | 29 | Y |
|  | 9 | M | 57 | 90 | L | 49 | 52 | 30 | 36 | 72 | 143 | 90 | 15 | 26 | N |
|  | 10 | M | 58 | 100 | L | 64 | 61 | 55 | 59 | 63 | 97 | 95 | 55 | 25 | N |
|  | 11 | M | 159 | 90 | L | 44 | 46 | 24 | 26 | 61 | 84 | 35 | 0 | 27 | N |
|  | 12 | M | 49 | 100 | L | 36 | 43 | 11 | 15 | 67 | 133 | 50 | 9 | 24 | N |
|  | 13 | M | 36 | 100 | R | 62 | 63 | 42 | 75 | 51 | 112 | 80 | 21 | 25 | Y |
|  | 14 | M | 29 | 100 | L | 61 | 63 | 44 | 49 | 52 | 91 | 50 | 59 | 20 | Y |
|  | 15 | M | 164 | 50 | R | 61 | 60 | 42 | 37 | 53 | 125 | 95 | 70 | ¥ | N |
|  | 16 | M | 48 | -70 | R | 50 | 48 | 25 | 36 | 99 | 154 | 100 | 76 | 26 | N |
|  | 17 | M | 69 | 100 | R | 49 | 52 | 36 | 37 | 66 | 105 | 35 | 56 | 24 | Y |
| Mean (SD) |  | 3 F / 14 M | 82.4 (58.4) | 67.4 (45.5) | 8 L / 9 R | 54.6 (10.6) | 54.4 (10.0) | 39.9 (17.2) | 42.2 (16.3) | 65.5 (13.6) | 115.8 (24.7) | 67.1 (30.6) | 40.1 (28.6) | 25.4 (2.4) | 6 Y / 11 N |
| Exercise |  |  |  |  |  |  |  |  |  |  |  |  |  |  |  |
|  | 1 | M | 141 | 60 | L | 63 | 63 | 100 | 95 | ^ | 116 | 60 | 21 | 26 | N |
|  | 2 | M | 138 | 100 | L | 57 | 56 | 43 | 44 | 57 | 113 | 90 | 27 | 27 | Y |
|  | 3 | M | 83 | 60 | R | 53 | 61 | 39 | 55 | 95 | 134 | 60 | 21 | 28 | N |
|  | 4 | M | 10 | 50 | L | 63 | 66 | 96 | ^ | 78 | 145 | 110 | 47 | 26 | N |
|  | 5 | M | 32 | 70 | L | 62 | 64 | 47 | 58 | 53 | 112 | 80 | 74 | 26 | N |
|  | 6 | F | 99 | 100 | L | 39 | 43 | 28 | 22 | 65 | 131 | 45 | 16 | 27 | N |
|  | 7* | F | 52 | 100 | L | 63 | 61 | 64 | 54 | 87 | 142 | 40 | 29 | 25 | N |
|  | 8 | M | 143 | 60 | R | 66 | ^ | 54 | 68 | 66 | 166 | 115 | 45 | 24 | Y |
|  | 9 | F | 104 | 58 | R | 57 | 58 | 54 | 57 | 56 | 123 | 55 | 36 | 26 | N |
|  | 10 | F | 95 | -75 | R | 23 | 19 | 14 | 21 | 58 | 134 | 70 | 63 | ^ | N |
|  | 11 | F | 114 | 100 | L | 62 | 59 | 43 | 42 | 97 | 126 | 35 | 56 | 27 | N |
|  | 12 | M | 51 | 80 | L | 65 | 64 | 55 | 54 | 65 | 139 | 145 | 107 | 26 | N |
|  | 13 | M | 29 | 60 | L | 62 | 64 | ^ | 81 | 90 | 151 | 135 | 24 | 23 | N |
|  | 14 | M | 80 | 100 | L | 25 | 30 | 15 | 24 | 61 | 146 | 95 | 82 | 30 | N |
| Mean (SD) |  | 5 F / 9 M | 83.6 (43.5) | 65.9 (45.0) | 10 L / 4 R | 54.3 (14.5) | 54.5 (14.7) | 50.1 (26.2) | 51.8 (22.0) | 71.4 (15.9) | 134.1 (15.5) | 81.1 (35.1) | 46.3 (27.1) | 26.2 (1.7) | 2 Y / 12 N |

### Post-intervention Motor Impairment and Function

Arm motor function improved for stroke participants as shown by faster rates of WMFT completion at the 24-hour retention test (main effect of Time *F*(1,27) = 4.312, *p* = .047, η_p_^2^ = .14). There were no changes in arm impairment as measured by the FMA.

### Serial Targeting Task: Motor Acquisition

Motor practice led to improved performance on the STT for both participant groups (Sequence × Practice Day interaction: *F*(4,276) = 5.284, *p* < .001, η_p_^2^ = .07); attributable to faster RTs for both random (*p*’s < .044) and repeated sequences (*p*’s < .001). We also found a Sequence × Practice Day × Participant Group interaction (*F*(4,276) = 3.688, *p* = .006, η_p_^2^ = .05). Post-hoc testing revealed this was driven by a Sequence × Practice Day interaction for the older adult group (*p* < .001) but not the stroke group (*p* = .745) (Figure 3).

**Figure 3.**
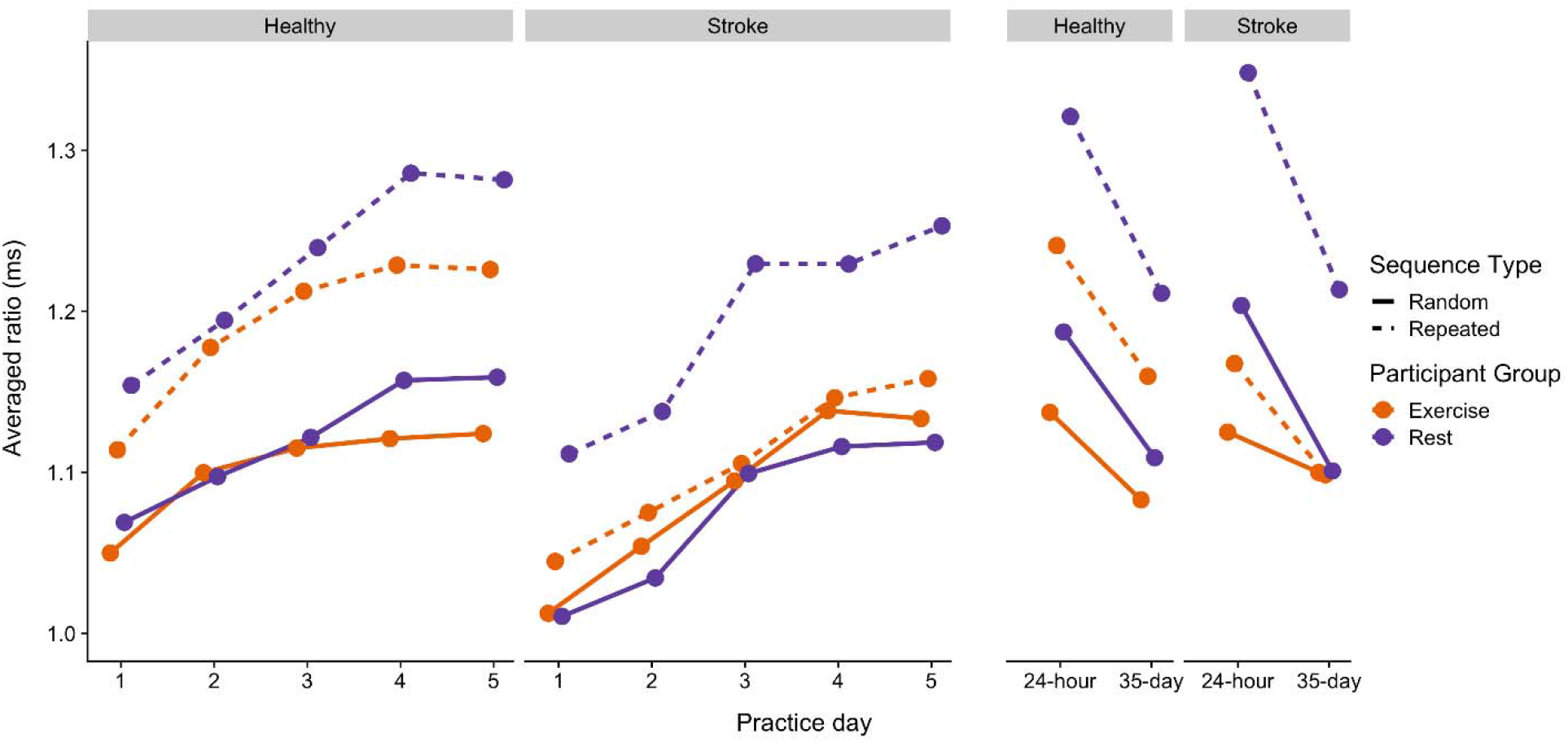
Averaged ratio RT as a function of practice day for stroke and healthy older adults for exercise (orange) and rest (purple). Values are averaged ratios relative to pre-test. Higher values indicate better motor performance (practice day 1-5) or motor learning (24-hour and 35-day retention).

### Serial Targeting Task: Retention Tests

Across both retention tests, we noted a Sequence × Intervention Group (*F*(1,64) = 5.650, *p* = .020, η_p_^2^ = .08) interaction. The rest (*p* < .001) but not exercise group (*p* = .054) showed a difference between the repeated and random sequences (Figure 3). The rest group also showed greater change in motor learning (main effect of Intervention Group *F*(1,64) = 5.959, *p* = .017, η_p_^2^ = .09) compared to the exercise group. Faster RTs were noted at the 24-hour compared to the 35-day retention test (main effect of Retention Day *F*(1,64) = 31.587, *p* < .001, η_p_^2^= .33). Finally, a main effect of Sequence (*F*(1,64) = 34.271, *p* < .001, η_p_^2^ = .35) revealed a greater change in repeated relative to the random sequences.

### Trail Making Test-A

A significant Participant Group × Intervention Group × Time interaction was observed (*F*(1, 66) = 5.204, *p* = .026, η_p_^2^ = .07). The stroke group took longer to complete the TMT-A as compared to older adults (main effect of Participant Group *F*(1, 66) = 6.723, *p* = .012, η_p_^2^ = .09), but the three-way interaction showed differences in how the participant groups responded to exercise. All older adults completed the TMT-A faster during post-intervention, however, the stroke exercise group completed the TMT-A faster than the rest group at the post-intervention session (Figure 4, Supplementary Figure 2).

**Figure 4.**
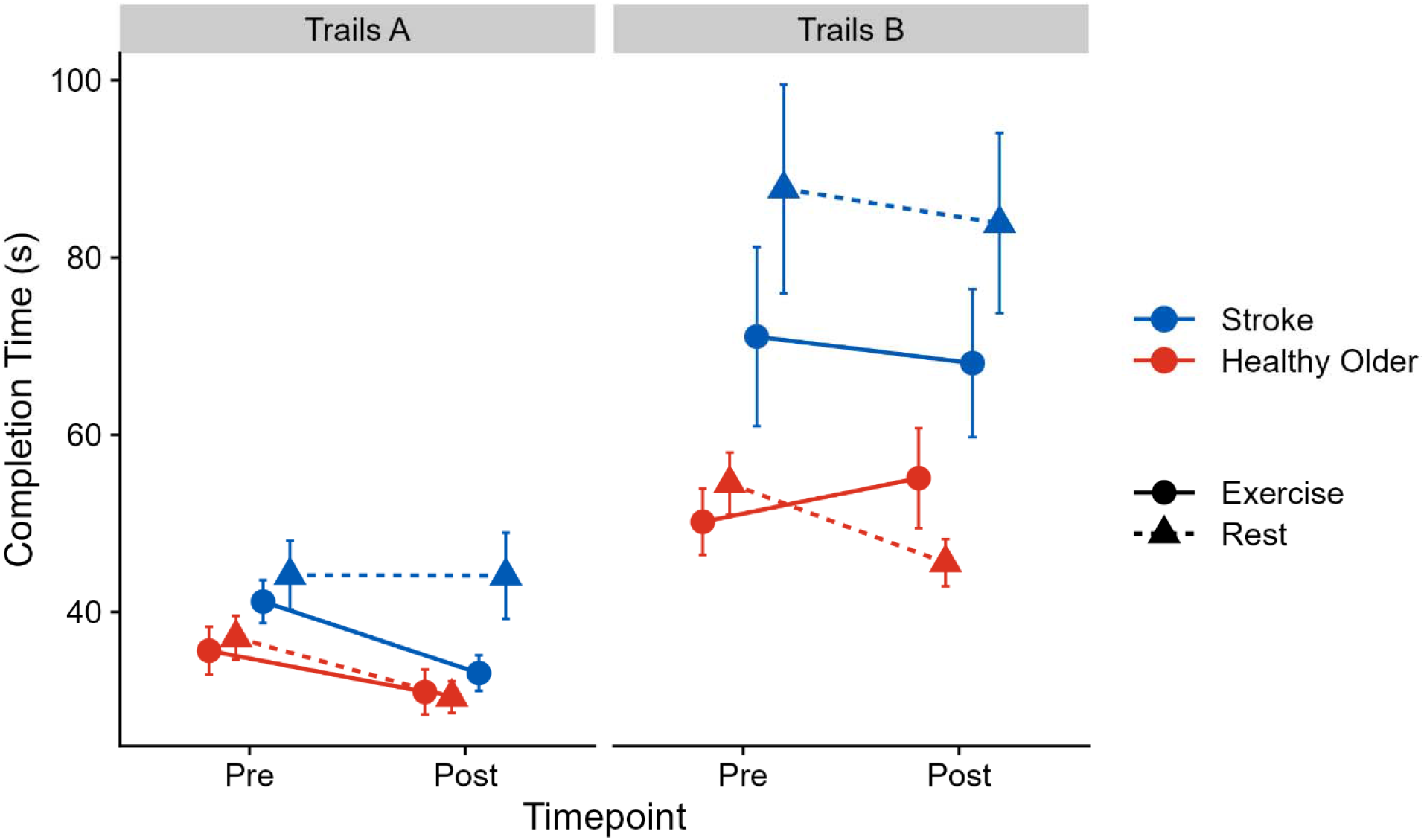
Mean total completion time (in seconds) for Trails Making Test-A (left) and Trails Making Test-B (right) disaggregated by exercise (solid line), rest (dashed lines), stroke (blue) and healthy older adults (blue) participants.

### Trail Making Test-B

There was a significant Intervention Group × Time interaction (*F*(1,66) = 4.506, *p* = .038, η_p_^2^ = .06) reflecting that TMT-B completion times improved with the rest but not the exercise group. A main effect of Participant Group was observed (*F*(1,66) = 17.616, *p* < .001, η_p_^2^ = .21), indicating stroke participants took longer to complete the TMT-B compared to older adults (Figure 4, Supplementary Figure 2).

### Object Hit and Avoid: Target Hits

Older adults hit more targets than stroke participants as evidenced by a main effect of Participant Group (*F*(1, 66) = 30.179, *p* < .001, η_p_^2^ = .31; Figure 5A). Participants hit more targets in the post-compared to pre-intervention, as evidenced by a significant main effect of Time (*F*(1, 66) = 15.361, *p* < .001, η_p_^2^ = .19), driven by improvements in the trained (i.e., affected/non-dominant hand) rather than untrained (i.e., the less-affected/dominant) hand whose performance did not change (Hand × Time interaction: *F*(1, 66) = 3.034, *p* = .086, η_p_^2^ = .04). Importantly, these changes were not at the expense of speed (see Supplementary Results). A significant Participant Group × Hand interaction (*F*(1, 66) = 28.140, *p* < .001, η_p_^2^ = .30) and main effect of Hand (*F*(1, 66) = 88.187, *p* < .001, η_p_^2^ = .57) demonstrates that the affected/non-dominant hand hit fewer targets than the less-affected/dominant hand, however this was driven by the stroke group as evidenced by the interaction. See Supplementary Figure 3,4 for individual differences.

**Figure 5.**
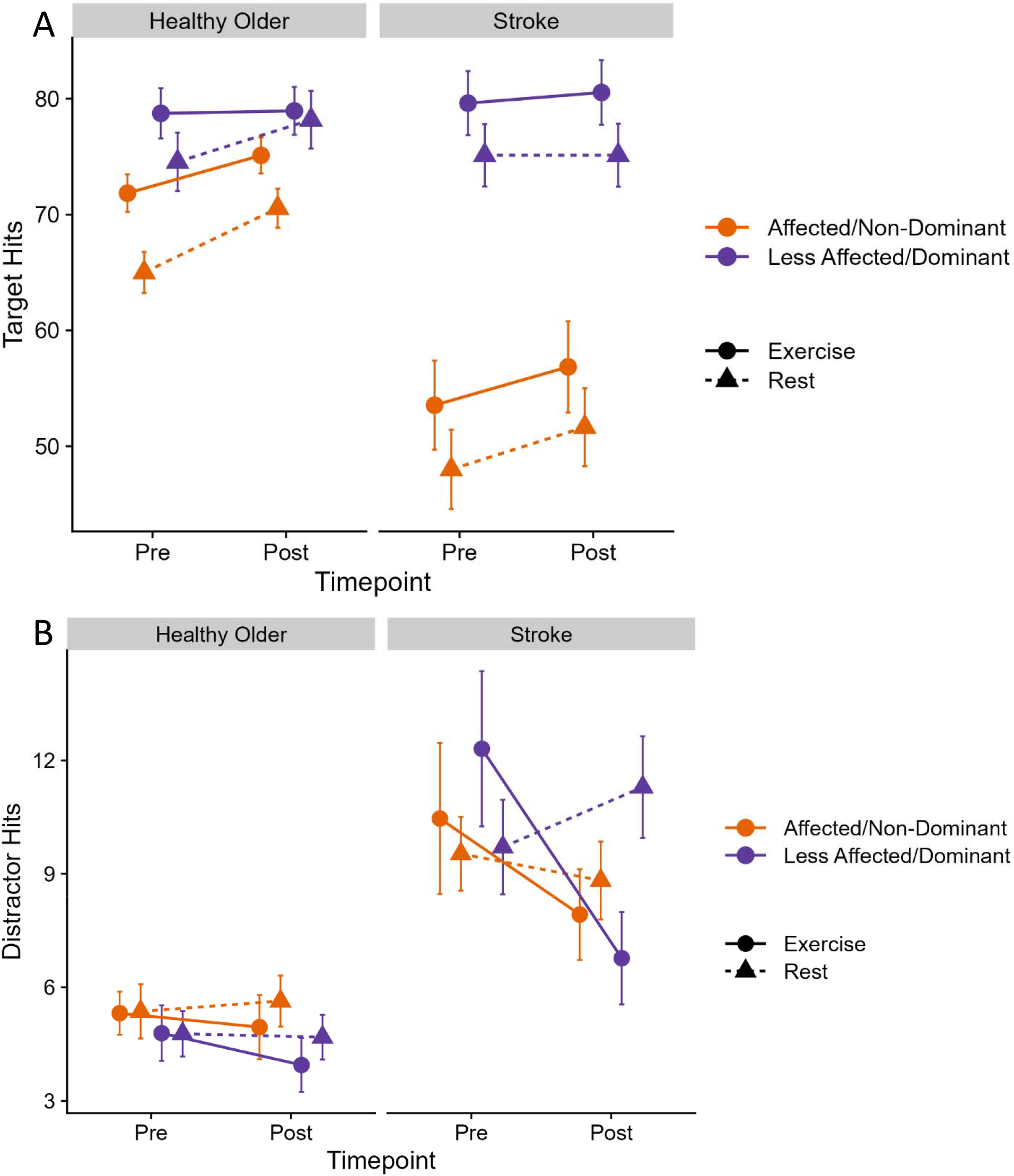
Mean number of A) target hits and B) distracter hits in object hit and avoid for healthy older adults (left) and stroke (right), disaggregated by exercise (solid lines), rest (dashed lines), and hand (orange: affected/non-dominant; purple: less affected/dominant).

### Object Hit and Avoid: Distractor Hits

A significant Intervention Group × Time interaction (*F*(1,66) = 4.613, *p* = .035, η_p_^2^ = .07) revealed that the number of distractors hit decreased over time for both stroke and older adults exercise groups (Figure 5B, Supplementary Figure 5,6). A significant Participant Group × Hand interaction (*F*(1,66) = 4.073, *p* = .048, η_p_^2^ = .06) demonstrates fewer distractor hits for the affected hand in the stroke group, while a main effect of Participant Group (*F*(1,66) = 27.418, *p* < .001, η_p_^2^ = .29) shows the stroke participants hit more distractors compared to older adults. There was also a Fitness z-score × Time interaction trend (*F*(1, 66) = 3.963, *p* = .051, η_p_^2^ =.06). A follow-up Spearman’s correlation revealed that those with higher baseline physical fitness showed greater gains (i.e., greater reduction in distractor target hits) from pre- to post-intervention, (ρ_s_ (71) = −.267, *p* = .025).

## Discussion

After a 5-day HIIT exercise or rest and motor practice intervention, all groups demonstrated motor acquisition and learning. There was no advantage of exercise for motor learning in either stroke participants or older adults, however those in the exercise group showed a benefit for unpracticed cognitive-motor tasks. Specifically, the stroke exercise group showed greater changes in processing speed (TMT-A) following the intervention. Participation in HIIT exercise paired with motor practice also led to improved visuospatial skills and inhibitory control (OHA) for both participant groups. Together, our findings suggest that exercise paired with motor task practice is a safe and effective means of enhancing cognitive-motor skills after stroke and in older adults.

Our findings suggest that multicomponent interventions that pair exercise with a task appear to improve motor tasks involving a high level of cognitive demand in individuals with stroke. For example, past work employing a single bout of high-intensity treadmill walking (Ploughman et al., 2008) or an 8-week exercise program (Quaney et al., 2009) improved upper-limb motor function, without affecting cognition. In hindsight, the selective improvement of motor function may be expected given that the intervention contained no cognitive engagement but was limited to exercise. In contrast, adaptive cognitive training improved cognition in individuals with chronic stroke when paired with either a 10-week aerobic exercise program or low activity range of motion exercises; however, the greatest cognitive gains were seen in the aerobic exercise group (Ploughman et al., 2019). Importantly, cognition did not improve with non-adaptive cognitive engagement, when paired with either aerobic exercise or low-demand physical activity (Ploughman et al., 2019). These data imply that cognitive training may confer the greatest benefits when paired with exercise in individuals with chronic stroke.

Adopting a multimodal approach, we paired exercise with motor practice. Interestingly, we found that exercise did not enhance motor learning of the STT beyond that induced by practice for both participant groups, but instead transferred to motor tasks that require selective cognitive capacities. Specifically, we observed enhanced processing speed (TMT-A) for stroke participants in the exercise group and enhanced visuomotor spatial skills and inhibitory control (OHA) in both participant groups who exercised before motor practice. These results are noteworthy because participants did not explicitly undergo any cognitive training during the intervention, but instead practiced an implicit motor sequence task following exercise or rest. These results suggest that exercise effects can transfer to unpracticed cognitive-motor tasks. Future studies testing the impact of pairing exercise with motor practice should also include cognitive assessments to understand whether this combination affects other cognitive functions.

Skill acquisition can play an instrumental role in enhancing cognitive function (Diamond & Ling, 2016). This idea stems from theories that posit that the early stages of motor learning place increased demands on cognition (Fitts & Posner, 1967). In addition, there is ample data showing links between exercise and improved cognitive function in older adults (Erickson et al., 2009, 2011; Voss et al., 2013). Here, we discovered that while both stroke and older adult groups demonstrated motor learning, only the exercise groups showed enhanced ability in cognitive-motor tasks. The impact of exercise plus motor learning transferred to improve processing speed, visuomotor skills, and inhibitory control required by the TMT-A and OHA tasks. To perform these tasks well, participants had to make small, distinct movements quickly and accurately while inhibiting anticipatory movements.

Processing and visuomotor speed, and visuoperceptual abilities are cognitive domains that are predictors of functional outcomes after stroke. In a large sample (n = 419), visuomotor speed but not attention, language, visuoconstruction, verbal or visual memory predicted post-stroke functional status (Narasimhalu et al., 2011). Similarly, only processing speed and visuoperceptual abilities predicted functional outcomes 5 years following a stroke (n = 307) whereas memory, executive function, and language did not (Barker-Collo et al., 2010). These results are relevent in the current study as we observed cognitive gains limited to processing speed (TMT-A) and visuomotor speed (OHA), but not executive function (TMT-B), suggesting that exercise paired with 5-days of motor practice selectively improve processing speed and visuoperceptual abilities, two of the most important cognitive measures in stroke.

There was no motor learning advantage for the exercise group. This is in contrast with at least one other study which found that stroke participants who first completed a single bout of HIIT exercise before motor practice displayed enhanced explicit motor learning when assessed the following day compared to a rest group (Nepveu et al., 2017). Here, we implemented an implicit motor sequence task. It is possible that exercise differentially affects implicit and explicit memory systems. Consistent with this hypothesis and our current findings, two studies using the same, implicit motor learning task also found no effect of exercise on skill acquisition in young adults (Baird et al., 2018; Mang et al., 2016). In contrast, young adults who exercised before practicing an explicit isometric pinch force sequencing task demonstrated better learning compared to those in the rest group (Statton et al., 2015). Similarly, participants who exercised before practicing a pursuit-rotor task (Tomporowski & Pendleton, 2018), repeatedly tracing a curve by flexion/extension arm movements (Thomas et al., 2016), or simple ballistic thumb acceleration movements (Opie & Semmler, 2019) also displayed superior learning compared to participants who rested. Collectively, exercise may preferentially affect explicit motor learning tasks where a cognitive rule is followed during learning. Future studies should directly test this hypothesis.

### Limitations

One limitation of the current study is that only HIIT exercise was tested. It is possible that other volumes, intensities, or frequencies of exercise would have a different effect. Also, we did not include a battery of other cognitive assessments. It is unknown if our findings are specific to motor tasks that require a type of cognitive control or if cognition in general was altered. Further, HIIT exercise may come with increased risks, especially for sedentary individuals with unknown underlying health conditions. However, all participants who were cleared for exercise in the current study were able to complete the HIIT exercise intervention without any adverse effects despite a wide range of physical fitness. With screening we found that our exercise protocol was feasible and can likely be generalized to other clinical populations.

### Conclusions

Five days of HIIT exercise paired with motor practice led to improved processing speed for individuals with stroke. Additionally, both exercise participant groups showed improved visuospatial skills and inhibitory control following our intervention. Together, exercise paired with motor practice appears to be a safe and effective means of enhancing cognitive-motor skills after stroke and in older adults.

## Data availability

All data will be available from the corresponding author upon reasonable request.

