## supplement for "Improved cognition after high-intensity exercise paired with motor practice in individuals with stroke and older adults: A randomized controlled trial"

**Methods**

*Explicit Awareness*

Testing of explicit awareness occurred during the 24-hour 30-35-day sessions. Each explicit awareness test involved showing participants a series of 10 sequence movies (in random order) displayed on the Kinarm workspace they had been practicing the serial targeting task on and asking them (yes/no) whether they recognized any of the sequences. Each movie consisted of 6 sequential targets: three of them were repeating sequences (having been seen during practice sessions) and seven were random sequences (novel and not having been seen before). Individuals who identified both the repeated and random sequences at a better than chance (i.e., 2 of 3 repeated sequences identified correctly and 4 of 7 novel, random sequences identified as not having been seen before), were considered to have gained explicit awareness.

*Statistical Analysis*

Baseline demographic data were evaluated using a 2 (Participant Group: stroke, controls) by 2 (Intervention Group: HITT exercise, rest) full factorial univariate ANOVAs using age, MoCA, and baseline fitness (composite Z-score). All significant main effects and interactions were followed by post-hoc pairwise comparisons, Sidak corrected.

**Results**

*Participant Dropout*

A total of 40 individuals in the chronic phase of stroke and 46 healthy older adults were consented to participate in this study (second middle box in CONSORT Figure 1 of the main body)**.** Among the stroke participants, 5 participants did not pass screening: 3 had their consent withdrawn despite being cleared for exercise, 1 was unable to be scheduled, and 1 did not meet eligibility criteria (they did not have adequate arm function to complete the STT). Three healthy older adult control participants did not pass screening: 2 participants were unable to be scheduled and 1 had an incidental finding during the stress test that was not cleared by a cardiologist. Therefore, 78 participants (stroke n = 35; healthy older adults = 43) were randomized to receive either HIIT exercise or rest. Eighteen stroke participants were allocated to the rest intervention: 2 discontinued the study. Specifically, 1 participant withdrew after health issues unrelated to the study, whereas 1 withdrew. It should also be noted that 2 stroke participants were lost to follow-up *after* the 24-hour retention session due to COVID-19 research closure, however, their motor practice and learning data (STT) and cognitive-motor data (pre- and post-intervention KST) were included in the final analyses. One stroke participant in the rest group was excluded from the final analysis because they could not follow the task instructions. Therefore, 17 stroke participants were included in the final analyses in the rest group. No healthy older adults discontinued, were lost to follow-up, or were excluded from the rest group. Therefore, 22 healthy older adult participants were included in the final analysis for the rest group. A total of 36 participants (stroke n = 15; controls n = 21) were randomized into the exercise group. No one discontinued the study or was lost to follow-up, however, 3 participants were excluded from the final analysis. One stroke participant and one healthy older adult (2 total) were excluded due to a data acquisition errors, whereas 1 healthy older adult participant was excluded because they could not follow the task instructions. Thus 14 stroke and 19 healthy older adult participants were included in the final analysis for the exercise group. See CONSORT Figure 1 in the main body of the manuscript for a summary. Also note that Kinarm Standard Test data (Trail Making Test-A, Trail Making Test-B, Object Hit and Avoid) from one participant from the exercise group (participant #7) was not collected and was therefore not included in the secondary analyses.

Demographics (stroke and older adults)

*Demographics: Age*

There was no main effect of Participant Group (*F* (1, 68) = .788, *p* = .378, η_p_^2^ = .01), Intervention Group (*F* (1, 68) = .081, *p* = .777, η_p_^2^ < .01), and no Participant Group by Intervention Group interaction (*F* (1, 68) = 1.562, *p* = .216, η_p_^2^ = .02) for age.

*Demographics: Baseline Fitness*

Likewise, there was no main effect of Participant Group (*F* (1, 68) = .212, *p* = .647, η_p_^2^ < .01), Intervention Group (*F* (1, 68) = .450, *p* = .505, η_p_^2^ = .01), and no Participant Group by Intervention Group interaction (*F* (1, 68) = .037, *p* = .849, η_p_^2^ < .01) for baseline fitness as measured by the z-composite score. Thus, both stroke and healthy older adult participant groups were of similar ages and had similar baseline physical fitness levels.

*Demographics: Montreal Cognitive Assessment (MOCA)*

For the analysis of MOCA score, a total of 6 scores were missing (stroke, n=27; healthy older adults, n=39). There was a main effect of Participant Group (*F* (1, 62) = 9.498, *p* = .003, η_p_^2^ = .13). Healthy older adults had overall higher MoCA scores (M = 27.41 ± 2.04) compared to the stroke group (M = 25.78 ± 2.10). There was no main effect of Intervention Group (*F* (1, 62) = .043, *p* = .836, η_p_^2^ < .01) and no Participant Group by Intervention Group interaction (*F* (1, 62) = 2.203, *p* = .143, η_p_^2^ = .03).

*Time between sessions* (stroke group)

There were no group differences in the number of days between practice sessions. The amount of time between each practice session was 3.9 days and the amount of time between the 24-hour and 35-day retention sessions was 33.6 days.

*Heart rate* (stroke group)

Heart rate (HR) was collected from 12 of the 14 stroke, exercise participants, with 10 complete datasets across the 5 days of the intervention. Average HR and RPE for each exercise period (rest, warm-up, HITT, recovery) can be found in Table 2.

A 5 (Day: 1, 2, 3, 4, 5) × 2 (Period: HITT, recovery) × 3 (Interval: 1, 2, 3) RM-ANOVA revealed a main effect of Day (*F*(4,36) = 2.693, *p* = .046, η_p_^2^ = .23), however post-hoc comparisons did not survive correction. There was also a main effect of Period (*F*(1,9) = 41.587, *p* < .001, η_p_^2^ = .82). Heart rate was significantly higher in the high-intensity Period (M = 114.94 ± 2.8) relative to the recovery Period (M = 99.19 ± 3.9) of exercise. Importantly, 75% of age-predicted max (220-average age of the exercise group) is 115.0, or .06 from averaged HIIT HR. There was also a main effect of Interval (*F*(2,18) = 9.140, *p* = .002, η_p_^2^ = .50). BPM were significantly higher in the third (M = 110.0 ± 3.4) compared to the first (M = 104.8 ± 3.0; *p* = .002) and second (M = 106.4 ± 3.2; *p* = .041) intervals. No interactions reached significance.

Serial Targeting Task

*Explicit Awareness (stroke group)*

There was no group difference in explicit awareness when assessed during the two retention intervals (24-h, 35-day). Individuals in the exercise group got 59% and 61% correct during the explicit awareness assessment during the 24-hour and 30-day retention sessions, respectively. Individuals in the rest group got 55% and 61% correct at the 24-hour and the 30-day retention sessions, respectively.

A total of 5 individuals were classified as being fully explicitly aware of the repeated implicit sequence (at least 2 out of 3 for the repeated sequence and at least 4 out of 7 for the foils), 4 of which were from the exercise group. Similarly, 5 individuals were explicitly aware of the implicit sequence, 4 of which were from the exercise group. Non-parametric tests revealed no statistical group differences.

*Object hit and avoid: Hand Speed*

There was no main effect of Participant Group (*F*(1, 67) = 878, *p* = .352, η_p_^2^ = .01), Intervention Group (*F*(1, 66) = .135, *p* = .715, η_p_^2^ < .01), or their interaction (*F*(1, 66) = .511, *p* = .477, η_p_^2^ = .01). Hand speed did not change pre- to post-intervention, as evidenced by the lack of any main effects of Time (*F*(1, 66) = .065, *p* = .800, η_p_^2^ < .01, Participant Group by Time (*F*(1, 67) = .185, *p* = .669, η_p_^2^ < .01), or Participant Group by Intervention Group by Time interactions (*F*(1, 66) = 2.555, *p* = .115, η_p_^2^ = .04).

There was evidence of a main effect of Hand (*F*(1, 66) = 43.849, *p* < .001, η_p_^2^ = .40) suggesting that the affected/non-dominant hand was slower than the less-affected hand. This effect was driven by the Stroke group, whose affected hand was significantly slower than the less affected hand as evidenced by the Participant Group by Hand interaction (*F*(1, 66) = 18.406, *p* < .001, η_p_^2^ = .22).

Hand effects did not vary with Exercise Group or Time as evidenced by the lack of a Hand by Time (*F*(1, 66) = .018, *p* = .893, η_p_^2^ < .01), Participant Group by Hand by Time (*F*(1, 66) = .250, *p* = .619, η_p_^2^ < .01), Intervention Group by Hand by Time (*F*(1, 67) = .022, *p* = .883, η_p_^2^ < .01), or Participant Group by Intervention Group by Hand by Time (*F*(1, 67) = .321, *p* = .573, η_p_^2^ = .01) interactions.

Table 1. Heart rate and rate of perceived exertion (RPE) averaged across the practice sessions separated by exercise period for the exercise, stroke group. Each practice session for the exercise group was 23 minutes in duration and consisted of a 5-minute warm-up, followed by three high-intensity intervals (HIIT) of exercise for 3 minutes interleaved with 3 active recovery periods for 3 minutes. Values are reported as means with standard deviation.

| **Period** | **Heart Rate** | **RPE** |
| --- | --- | --- |
| Resting | 77.2 ± 13.4 |  |
| Warm-up | 91.4 ± 14.8 | 2.3 ± 1.2 |
| HITT 1 | 110.4 ± 13.9 | 4.9 ± 1.2 |
| Recovery 1 | 99.9 ± 14.7 | 2.7 ± 1.3 |
| HITT 2 | 115.7 ± 14.2 | 5.5 ± 1.4 |
| Recovery 2 | 100.6 ± 14.1 | 3.0 ± 1.4 |
| HITT 3 | 119.6 ± 14.6 | 5.9 ± 1.5 |
| Recovery 3 | 102.6 ± 14.0 | 2.9 ± 1.6 |

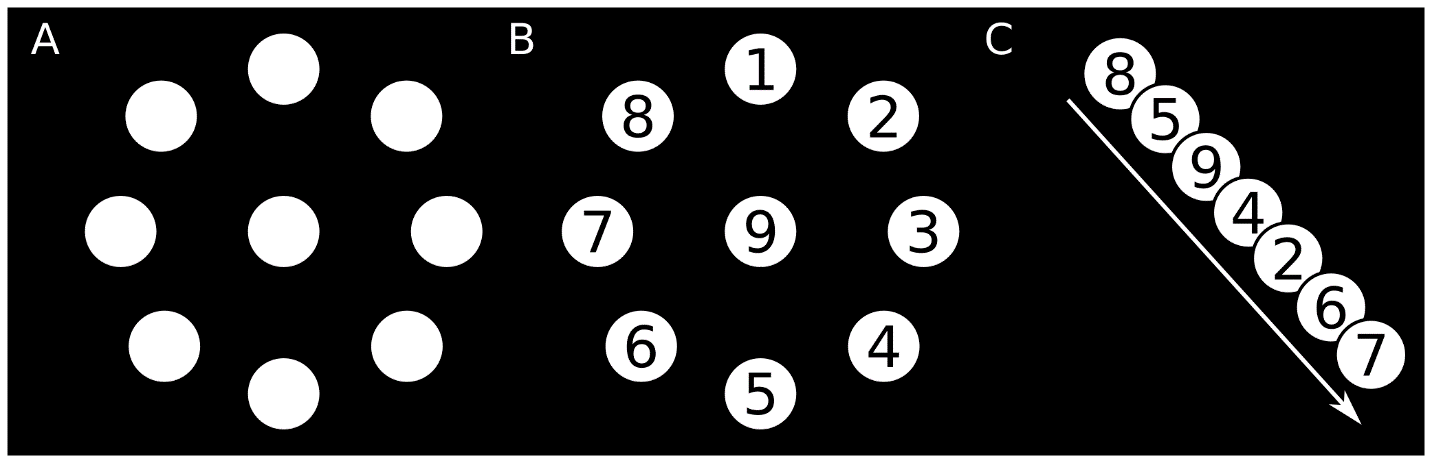

Figure 1. Spatial location and order of repeated targets in the motor task. A & B) Displays the spatial location of all possible targets. Note, targets were *not* numbered during the task. C) Order of the repeated sequence.

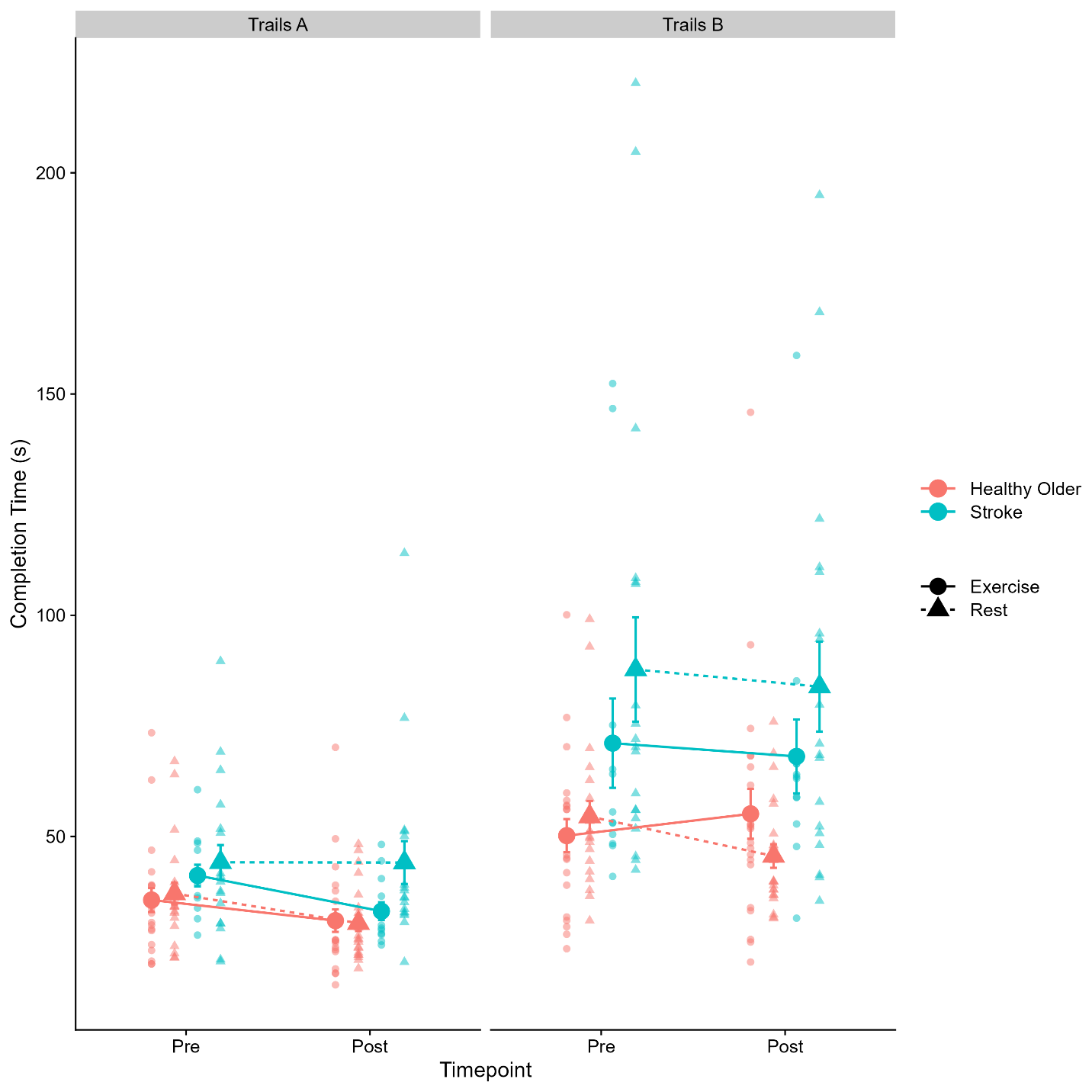

Figure 2. Group means (large circles and triangles) and individual (small circles and triangles shapes) of total completion time (in seconds) for Trail Making Test-A (left) and Trail Making Test-B (right) disaggregated by healthy older adults (pink) and stroke (blue), exercise (solid lines) and rest (dashed lines) groups.

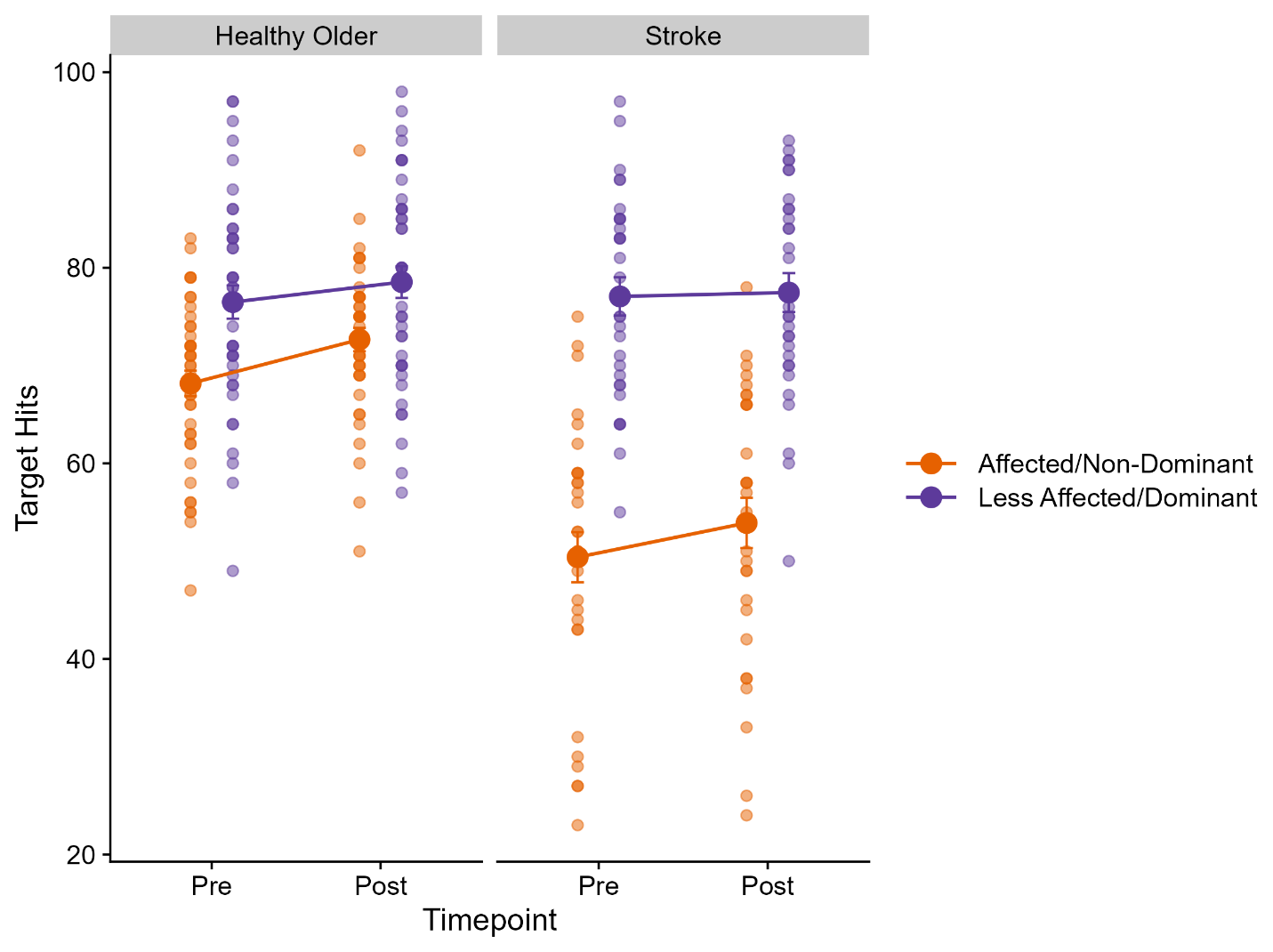

Figure 3. Group means (large circles) and individuals (small circles) number of targets hit in the object hit and avoid Kinarm task separated by affected / non-dominant (orange circles) or less affected / dominant (purple circles) for healthy older adults (left) and stroke (right) groups. Note, Data is collapsed across exercise and rest groups.

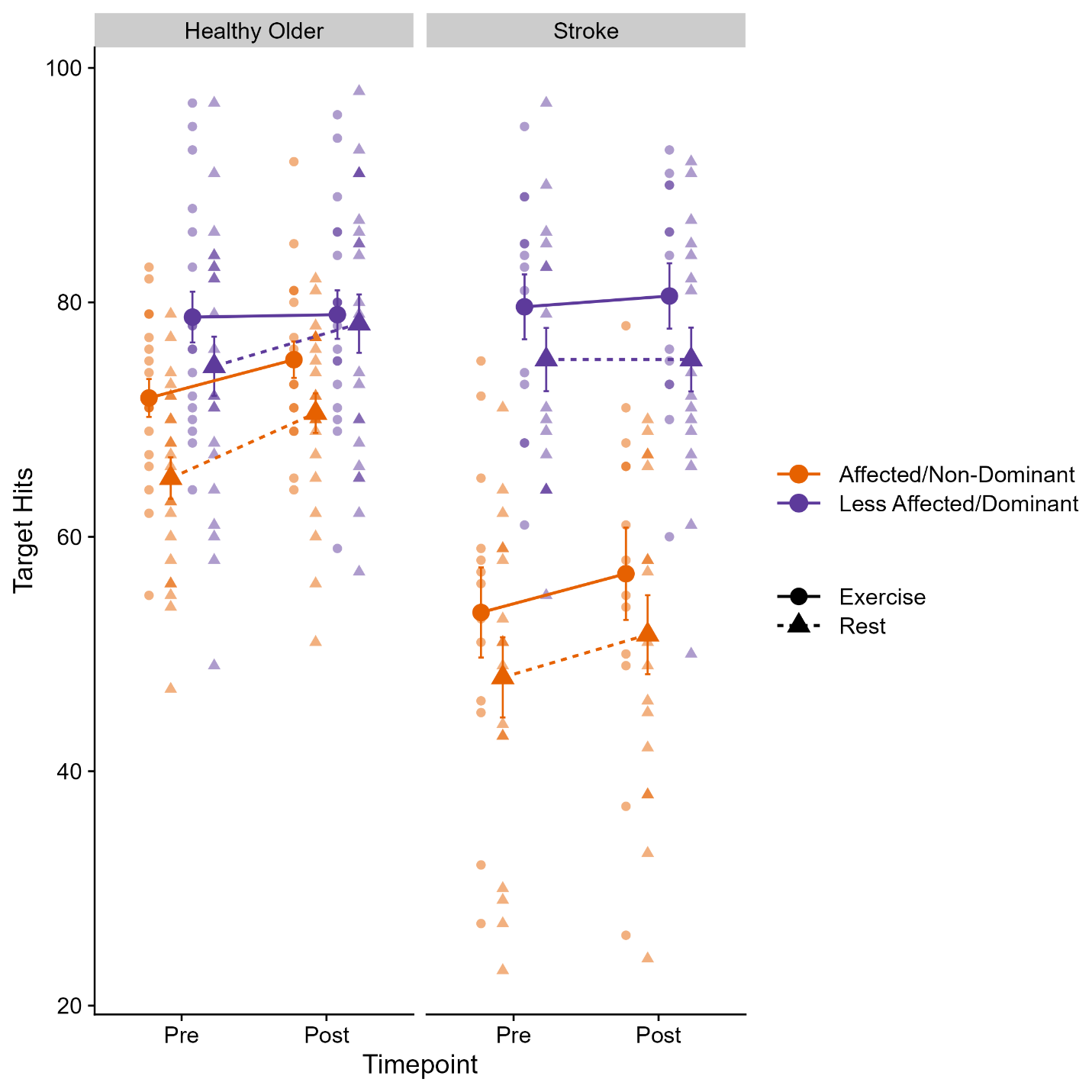

Figure 4. Group means (large circles and triangles) and individual (small circles and triangles) number of target hits in the object hit and avoid Kinarm task disaggregated by healthy older adults (left) stroke (right), exercise (circles), rest (triangles), affected / non-dominant hand (orange), and less affected / dominant (purple).

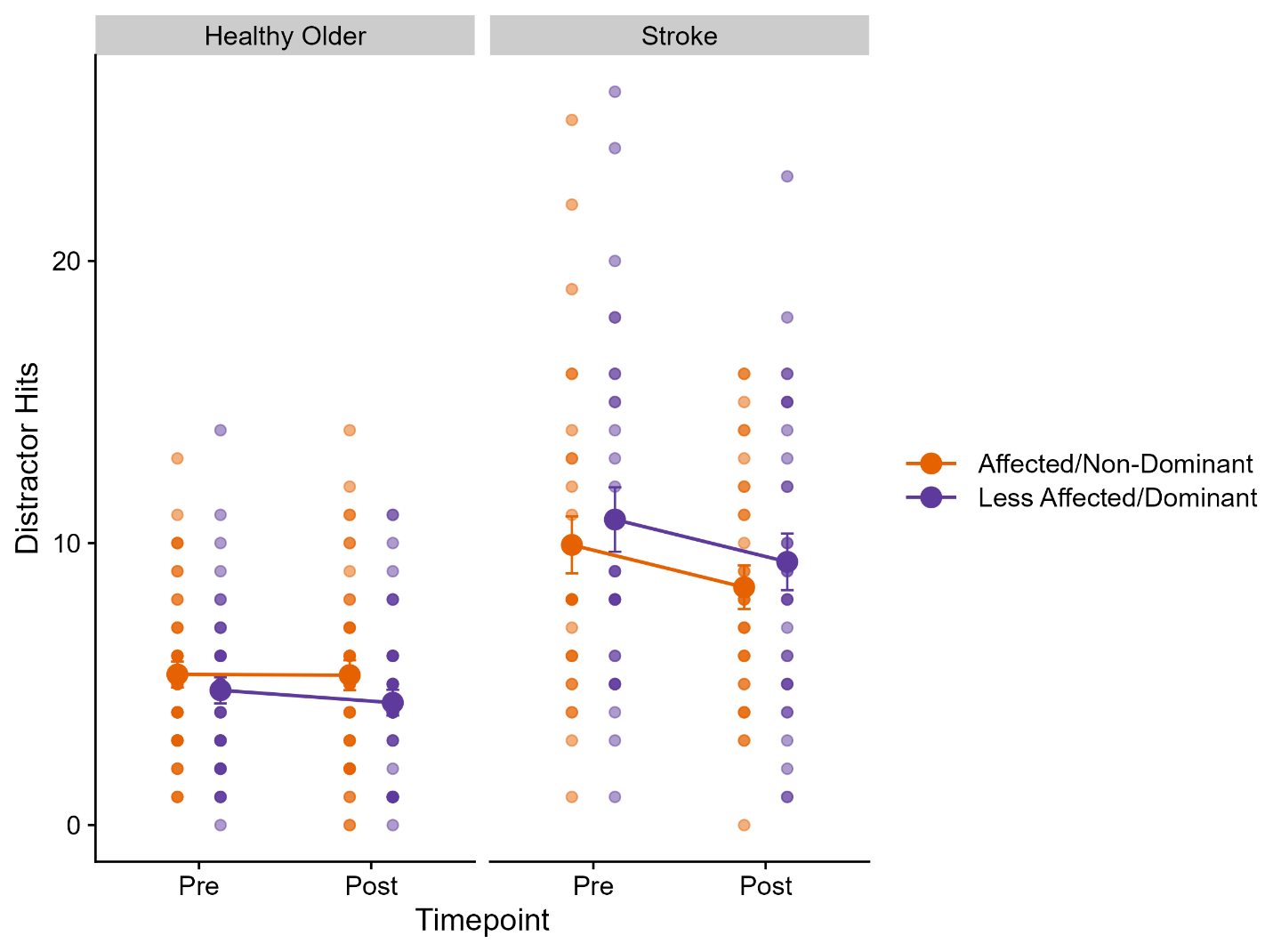

Figure 5. Group mean (large circles) and individual (small circles) number of distractors hit in the object hit and avoid Kinarm task collapsed for healthy older adults (left), stroke (right), disaggregated by affected / non-dominant hand (orange circles) and less affected / dominant (purple circles) hand. Note, data is collapsed across exercise and rest groups.

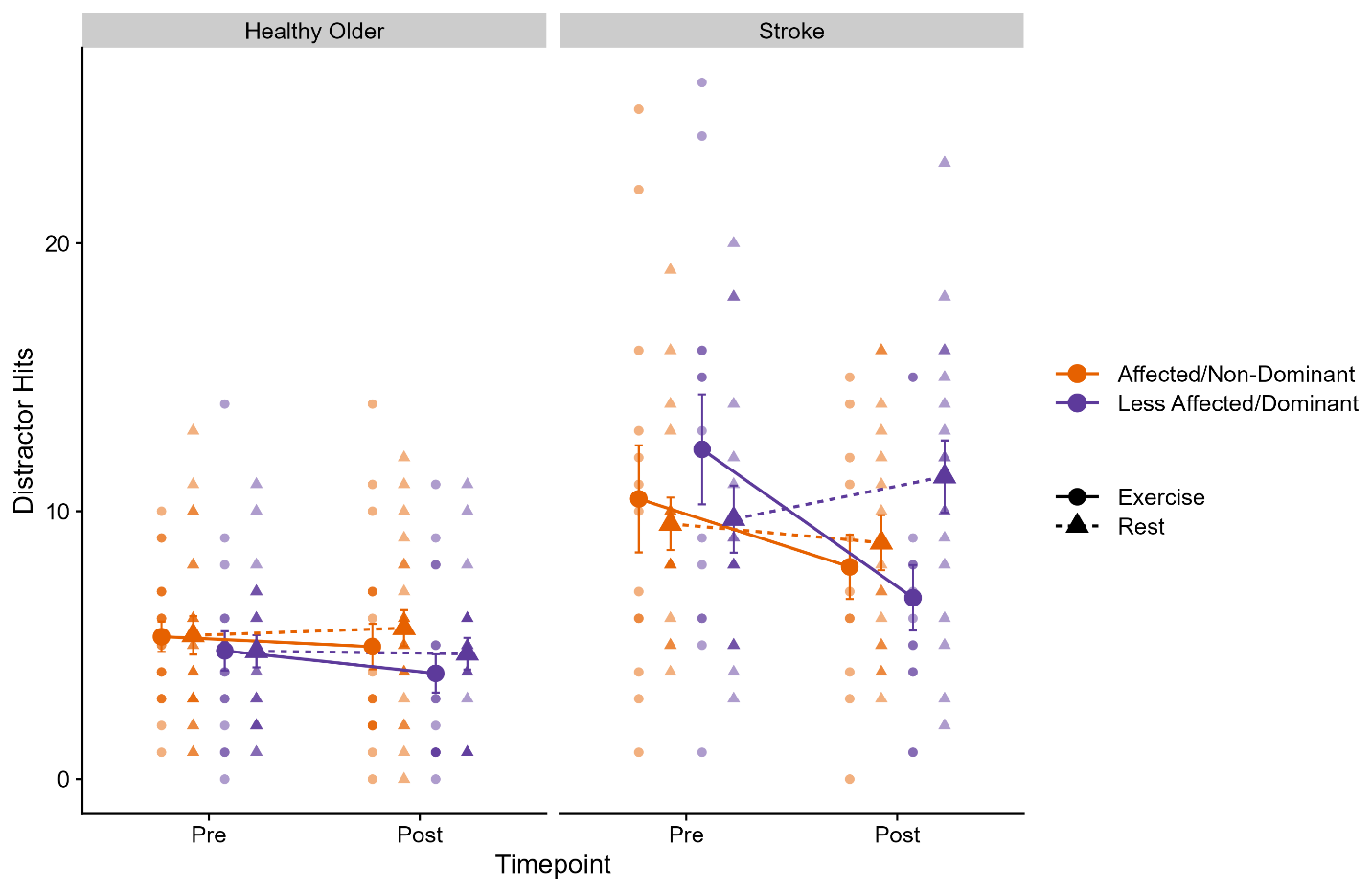

Figure 6. Group means (large circles) and individual (small circles) number of distractors hit in the object hit and avoid task collapsed for healthy older adults (left), stroke (right), disaggregated by exercise (circles) and rest (triangles), affected / non-dominant hand (orange) and less affected / dominant (purple) hands.
